# Bias and Variance of Adjusting for Instruments

**DOI:** 10.64898/2026.03.13.26348328

**Authors:** George Hripcsak, Tara Anand, Hsin Yi Chen, Linying Zhang, Yong Chen, Marc A. Suchard, Patrick B. Ryan, Martijn J. Schuemie

## Abstract

Propensity score adjustment is commonly used in observational research to address confounding, but controversy persists about how to select potential confounders. A desire to include all possible confounders is offset by a concern that more covariates will worsen bias or variance. Adjusting for an instrument, which affects the treatment but not the outcome, can increase bias due to unadjusted confounding and increase the variance of the effect estimate. Studies have shown that instruments may have a limited effect in practice, but a strong instrument can have a strong effect.

Diagnostics can bound the effect of instruments. Large-scale propensity score (LSPS) adjustment employs two diagnostics that assess the combined strength of instruments and confounders, ceasing the analysis if any covariate exceeds a correlation coefficient of 0.5 with the treatment and checking for an aggregation of instruments with equipoise reported as a preference score.

We assessed the impact of adjusting for instruments in the context of these diagnostics. In our simulations, which included single and multiple instruments and unmeasured confounding, when the diagnostics were satisfied, the increase in bias and variance due to instruments was less than the original bias and variance. This held true even when the instrument was 20 times stronger than the confounder in terms of variance contributed to the treatment. Our simulation aligns well with a previous assessment of the impact of adjusting for instruments and with separate empirical evidence that adjusting for many covariates surpasses attempts to identify a limited set of confounders.

## INTRODUCTION

One of the common approaches to addressing confounding in observational research is to use propensity score adjustment [1] in a comparative cohort study. There has been a decades-long argument about what covariates to include in the propensity model as potential confounders, contrasting inclusion of all pre-treatment covariates [2-4] with careful selection of confounders [5-8]. Instruments, which are covariates that are associated with the treatment but not the outcome, have been identified as one of the risks of too-broad inclusion: adjusting for an instrument always increases the bias of the effect estimate in the setting of unadjusted confounding if the model is linear [9]. In a complex field like medicine, some degree of residual unmeasured confounding remains likely, increasing concern about instruments. Adjusting for instruments also increases the variance of the estimate [10-12] even without unmeasured confounding.

A number of pieces have been published about the phenomenon [13-15]. A broader theoretical treatment explored the conditions under which a non-linear model is subject to bias amplification [16]. One study showed that for so-called “near-instruments,” which are confounders with a relatively weak association with the outcome, the benefit of adjusting for the confounder outweighs the cost of it being nearly an instrument [17]. A subsequent piece argued that the relative risk of instruments increases as the number of instruments and confounders increases [18]: the effect of instruments is additive, always pulling away from the true effect, but the effect of confounders is not because some may pull the estimate away from the true value and some may pull it in the opposite direction. Of course, instruments are included as a byproduct of trying to capture as many confounders as possible, so the end result depends on how well confounders are covered, which is an empirical question.

Current practice in confounder selection varies. Some guidelines assert that the number of variables adjusted for should be limited [19], with researchers often relying on domain experts to identify a small set of potential confounders. A larger set is selected by a technique called High-Dimensional Propensity Score (hdPS) adjustment, which employs the empirical selection of potential confounders based on the association with the treatment, the association with the outcome, and the prevalence [20], and it generally includes around 200 covariates. While in theory, hdPS should avoid instruments, given the very low prevalence of outcomes in most medical safety studies and many medical effectiveness studies, the ability to stably find all confounders from the tens of thousands of available covariates is limited.

A technique called Large-Scale Propensity Score (LSPS) adjustment [21,22] includes all pre-treatment covariates with some exceptions noted below. Repeated studies on real-world data have shown that this approach, which includes numerous covariates, to be superior to manual confounder selection [23,24] and to hdPS [21], and LSPS has also been shown to have emergent properties adjusting for unmeasured confounders indirectly [22]. A key component of LSPS is its diagnostics [22,25]. First, the analysis is rejected if any covariate has a Pearson correlation coefficient of 0.5 or greater with the treatment, and second, the equipoise of the propensity scores of the treatment groups is quantified with Walker’s preference score [26], which transforms the propensity score to reveal the overall ability of included covariates to predict treatment assignment. These diagnostics, which cannot separate the effect of instruments from confounders, nevertheless bound the potential strength of instruments. Variables with too-high correlation must either be identified as instruments by domain knowledge and removed from the propensity model or the hypothesis must be altered (e.g., if two medications are always given together then the hypothesis includes the effects of both drugs). The preference score diagnostic ensures that even if there is no single strong instrument that no combination of covariates is strongly correlated with the treatment. The question still remains what is the potential effect of instruments that are weak enough to pass diagnostics and remain in the propensity model.

A recent study of instruments in propensity score adjustment looked at the impact of including instruments in the propensity model using simulation and real-world data [27]. They found that instruments had minimal impact on the effect estimate and that using all covariates produced the best estimate. In addition, for a covariate that they selected as an instrument—study year— they found that it was a likely confounder. That is, even the study authors had difficulty separating confounders from instruments, reflecting the difficulty of this task and the danger of assuming confounders can be manually identified.

Despite the wealth of previous work, the operating characteristics of propensity score adjustment that includes an instrument and unadjusted confounding remains poorly quantified: what level of instrument strength causes what level of bias? In this study, we use simulation to assess the effect of instruments on bias and variance in estimation studies. We employ both LSPS diagnostics to see how the effect on bias and variance changes with different levels of the diagnostics. We begin with the nominal LSPS thresholds but explore a range of diagnostic thresholds.

## METHODS

The goal of our simulation was to assess the effect of adjusting for an instrument in the setting where there was unadjusted confounding. We varied the strength of the instrument and compared the bias and variance of a series of logistic regression models. Because our logistic model is non-collapsible, we included a second unadjusted instrument to keep the total variance due to instruments constant and therefore keep the effect of the confounder constant. This best reflected a real-world data experiment in which the variance is already set and the decision is whether to adjust for a variable.

In our simulation, we had confounder *X*, measured instrument *Z*, unmeasured instrument *U*, treatment *T*, and outcome *Y* defined as follows:

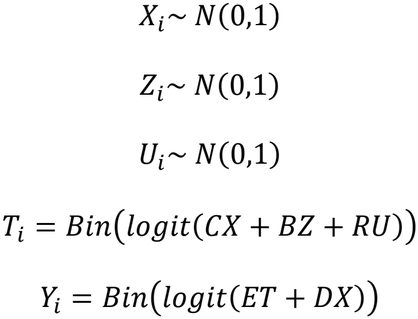

where C and D set the strength of confounding, B sets the strength of a measured instrument, R sets the strength of an unmeasured instrument, and E sets the strength of the effect of treatment on the outcome. We varied the strength of the measured instrument keeping the total variance of the treatment constant by also adjusting the strength of the unmeasured instrument such that *B*^2^ + *R*^2^ was constant.

We ran several logistic regression models for each set of simulation parameters using R’s glm function with family set to binomial. A crude model (M_crude_) included no covariates. Other models included just the instrument (M_instr_), just the confounder (M_conf_), and both the confounder and the instrument (M_conf-instr_). The model that includes only the confounder should asymptotically produce a proper estimate of the treatment effect on the outcome. The model that includes only the instrument illustrates the effect adjusting for an instrument in the setting of unmeasured confounding. The model that includes both the confounder and the instrument illustrates the effect adjusting for an instrument when all confounding is adjusted for. All simulations used a sample size of 200,000. We compared bias in the treatment effect estimate and variance of the treatment effect estimate. We also calculated the Pearson correlation coefficient of the adjusted instrument with the treatment to mirror the LSPS diagnostic, and we calculated equipoise [28] from propensity scores estimated by a logistic regression of instrument to predict the treatment (the confounder was not included to illustrate the case where there is unmeasured confounding).

We chose *E*=0.5, *C*=1, *D*=1, *B* in (1, 2, …, 7), and *B*^2^ + *R*^2^ = 50. We also varied B until we found the value that produced a correlation coefficient near 0.5, which is LSPS’s threshold for rejecting the analysis.

We repeated the analysis using 100 independent instruments each distributed as *N*(0,1) and sharing the same coefficient, *B*. We chose *E*=0.5, *C*=4, *D*=1, *B* from 1 to 3 by 0.25, and 100*B*^2^ + *R*^2^ = 1000.

We created a separate simulation of longitudinal time-to-event data and used Cox proportional hazards regression to analyze the data. We set treatment assignment as above with 100 instruments. We set the outcome hazard as follows:

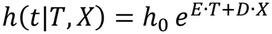

where here, *E* was the log hazard ratio for treatment, *D* was the log hazard ratio for the confounder, and *h*_0_ was the baseline hazard, set at 0.1. Censoring was independent of outcome and had censoring hazard rate of 0.02. We generated time to event or censoring, capping the follow up to 5 time units. We analyzed the data using R coxph for the Cox proportional hazards regression.

## RESULTS

All simulations ran to completion. Table 1 shows the single-instrument runs in detail. In Figure 1, we see the effect of adjusting for an instrument in the setting of unadjusted confounding. The estimate obtained by adjusting for just the confounder closely matches the simulation ground truth of 0.5. The crude estimate with no adjustment hovers at 0.6 due to confounding bias. The estimate obtained by adjusting for just the instrument deviates from 0.6 increasingly as the instrument strength increases. At an instrument strength of 4.65, the instrument has a Pearson correlation coefficient just over 0.5 with the treatment and equipoise just under 0.5, representing a study that would narrowly fail both LSPS’s diagnostic thresholds. This point is plotted on the graph and increases the effect estimate to 0.65, adding only 50% to the existing confounding bias despite failing both thresholds. When both the instrument and the confounder are adjusted for, then the estimate is unbiased at 0.5.

**Table 1.** Effect estimate and variance by adjustment for single instrument.

| **Treatment effect (E)** | **Confounder to treatment (C)** | **Confounder to outcome (D)** | **Adjusted instrument to treatment (B)** | **Unmeasured instrument to treatment (R)** | **Crude** |  | **Adjust instrument** |  | **Adjust confounder** |  | **Adjust instrument and confounder** |  | **Correlation treatment with adjusted instrument** | **Equipoise** |
|---|---|---|---|---|---|---|---|---|---|---|---|---|---|---|
|  |  |  |  |  | **Estimate** | **Variance** | **Estimate** | **Variance** | **Estimate** | **Variance** | **Estimate** | **Variance** |  |  |
| 0.5 | 1 | 1 | 1 | 7.000 | 0.596 | 0.000083 | 0.598 | 0.000084 | 0.497 | 0.000100 | 0.497 | 0.000101 | 0.108 | 1.000 |
| 0.5 | 1 | 1 | 2 | 6.782 | 0.601 | 0.000083 | 0.608 | 0.000087 | 0.508 | 0.000100 | 0.507 | 0.000105 | 0.218 | 0.935 |
| 0.5 | 1 | 1 | 3 | 6.403 | 0.592 | 0.000083 | 0.608 | 0.000093 | 0.502 | 0.000099 | 0.498 | 0.000112 | 0.327 | 0.752 |
| 0.5 | 1 | 1 | 4 | 5.831 | 0.609 | 0.000083 | 0.654 | 0.000102 | 0.514 | 0.000099 | 0.514 | 0.000123 | 0.434 | 0.570 |
| 0.5 | 1 | 1 | 4.65 | 5.327 | 0.593 | 0.000083 | 0.648 | 0.000112 | 0.496 | 0.000100 | 0.485 | 0.000135 | 0.507 | 0.461 |
| 0.5 | 1 | 1 | 5 | 5.000 | 0.599 | 0.000083 | 0.676 | 0.000118 | 0.507 | 0.000100 | 0.508 | 0.000142 | 0.543 | 0.410 |
| 0.5 | 1 | 1 | 6 | 3.742 | 0.603 | 0.000083 | 0.736 | 0.000145 | 0.499 | 0.000100 | 0.492 | 0.000175 | 0.650 | 0.272 |
| 0.5 | 1 | 1 | 7 | 1.000 | 0.597 | 0.000083 | 0.840 | 0.000197 | 0.510 | 0.000099 | 0.516 | 0.000239 | 0.758 | 0.123 |

**Figure 1.**
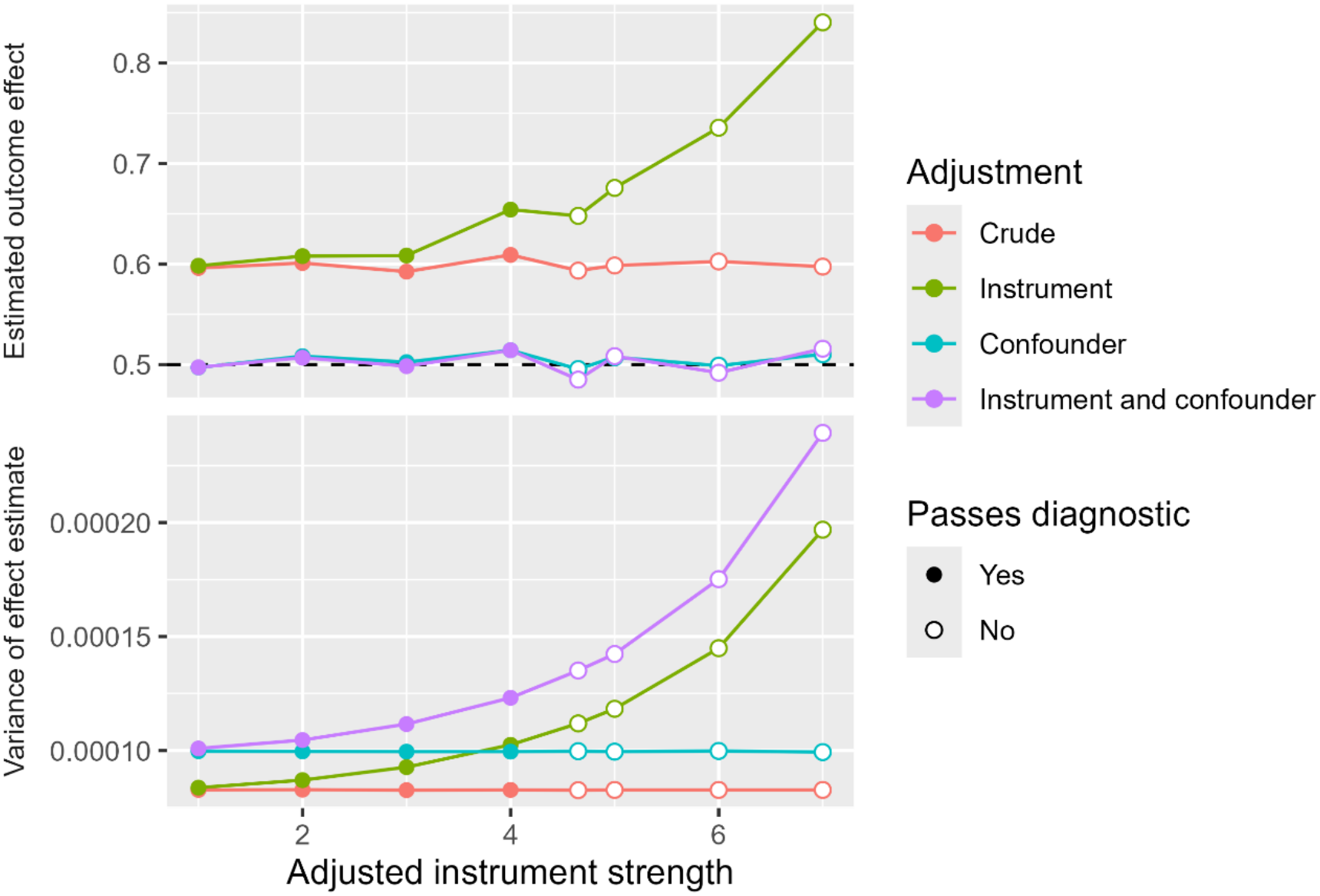
Effect of adjusting for a single instrument on estimation. The estimated effect of the treatment on the outcome is shown in the top graph with the correct effect shown as a dashed line at 0.5. Four adjustment methods are shown for a range of adjusted instrument strengths. Analyses that pass the LSPS diagnostics are shown as solid discs, and those that fail are shown as open circles. The bottom graph shows the variance of the effect estimate for the employed analyses and adjusted instrument strengths.

The variance of the estimate is also shown in Figure 1. The crude (biased) estimate has the lowest variance at 0.000083. Adjusting for just the confounder increases the variance to 0.000100. Adjusting for just the instrument increases the variance with increasing instrument strength. At 4.65, the increase is to 0.000112, again less than a 50% increase. Adjusting for both the confounder and the instrument leads to an increase that is more than additive but still less than double the crude variance at 4.65.

Table 2 and Figure 2 show the effect of adjusting for 100 instruments. For runs in which equipoise exceeded 0.5 and even down to 0.473, effect estimate bias never rose over the bias due to confounding, and the excess variance never exceeded 50% of the base variance. The per-instrument correlation with treatment was 0.050 at an equipoise of 0.473, illustrating the ability of equipoise to reveal the aggregate effect of multiple instruments. (We repeated the analysis for 10 instead of 100 instruments, and the results were nearly identical.) Table 3 and Figure 3 show the effect of adjusting for 100 instruments in the context of a Cox proportional hazards regression on longitudinal time-to-event data. For runs in which equipoise exceeded 0.5, effect estimate bias never rose over the bias due to confounding, and the excess variance never exceeded 50% of the base variance.

**Table 2.** Effect estimate and variance by adjustment for 100 instruments.

| **Treatment effect (E)** | **Confounder to treatment (C)** | **Confounder to outcome (D)** | **Adjusted instruments to treatment (B)** | **Unmeasured instrument to treatment (R)** | **Crude** |  | **Adjust instruments** |  | **Adjust confounder** |  | **Adjust instruments and confounder** |  | **Correlation treatment with each adjusted instrument** | **Equipoise** |
|---|---|---|---|---|---|---|---|---|---|---|---|---|---|---|
|  |  |  |  |  | **Estimate** | **Variance** | **Estimate** | **Variance** | **Estimate** | **Variance** | **Estimate** | **Variance** |  |  |
| 0.5 | 4 | 1 | 1 | 30.000 | 0.587 | 0.0000827 | 0.603 | 0.0000883 | 0.508 | 0.0000993 | 0.511 | 0.0001060 | 0.023 | 0.894 |
| 0.5 | 4 | 1 | 1.25 | 29.047 | 0.592 | 0.0000827 | 0.608 | 0.0000918 | 0.503 | 0.0000995 | 0.500 | 0.0001105 | 0.031 | 0.781 |
| 0.5 | 4 | 1 | 1.5 | 27.839 | 0.576 | 0.0000825 | 0.609 | 0.0000969 | 0.498 | 0.0000995 | 0.503 | 0.0001167 | 0.035 | 0.659 |
| 0.5 | 4 | 1 | 1.75 | 26.339 | 0.584 | 0.0000825 | 0.620 | 0.0001027 | 0.502 | 0.0000990 | 0.497 | 0.0001231 | 0.040 | 0.563 |
| 0.5 | 4 | 1 | 2 | 24.495 | 0.584 | 0.0000826 | 0.639 | 0.0001107 | 0.502 | 0.0000995 | 0.500 | 0.0001332 | 0.050 | 0.473 |
| 0.5 | 4 | 1 | 2.25 | 22.220 | 0.573 | 0.0000825 | 0.649 | 0.0001219 | 0.491 | 0.0000995 | 0.488 | 0.0001470 | 0.055 | 0.382 |
| 0.5 | 4 | 1 | 2.5 | 19.365 | 0.570 | 0.0000826 | 0.689 | 0.0001373 | 0.485 | 0.0000993 | 0.499 | 0.0001652 | 0.064 | 0.302 |
| 0.5 | 4 | 1 | 2.75 | 15.612 | 0.592 | 0.0000827 | 0.736 | 0.0001590 | 0.511 | 0.0000995 | 0.505 | 0.0001918 | 0.067 | 0.224 |
| 0.5 | 4 | 1 | 3 | 10.000 | 0.573 | 0.0000826 | 0.802 | 0.0001917 | 0.488 | 0.0000995 | 0.507 | 0.0002332 | 0.076 | 0.137 |

**Table 3.**
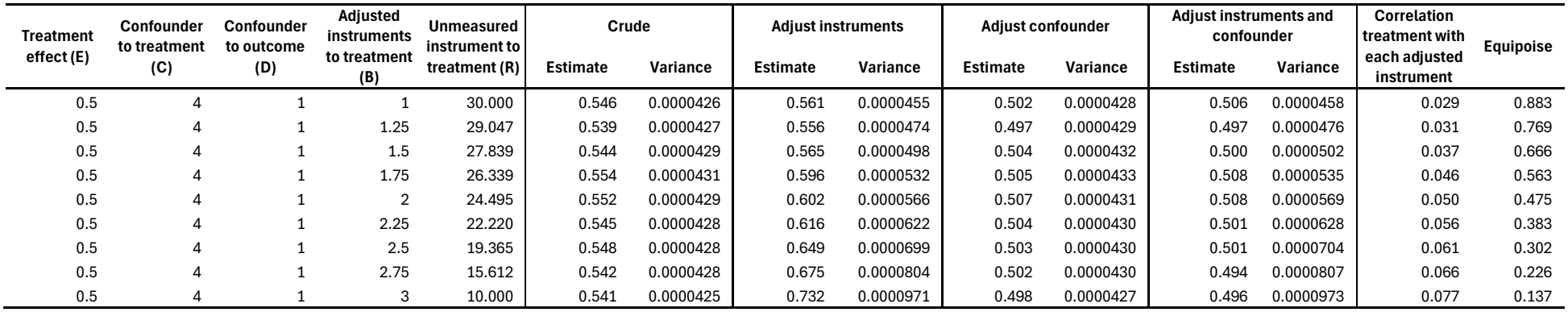
Effect estimate and variance by adjustment for 100 instruments using a Cox proportional hazards regression on longitudinal time-to-event data.

**Figure 2.**
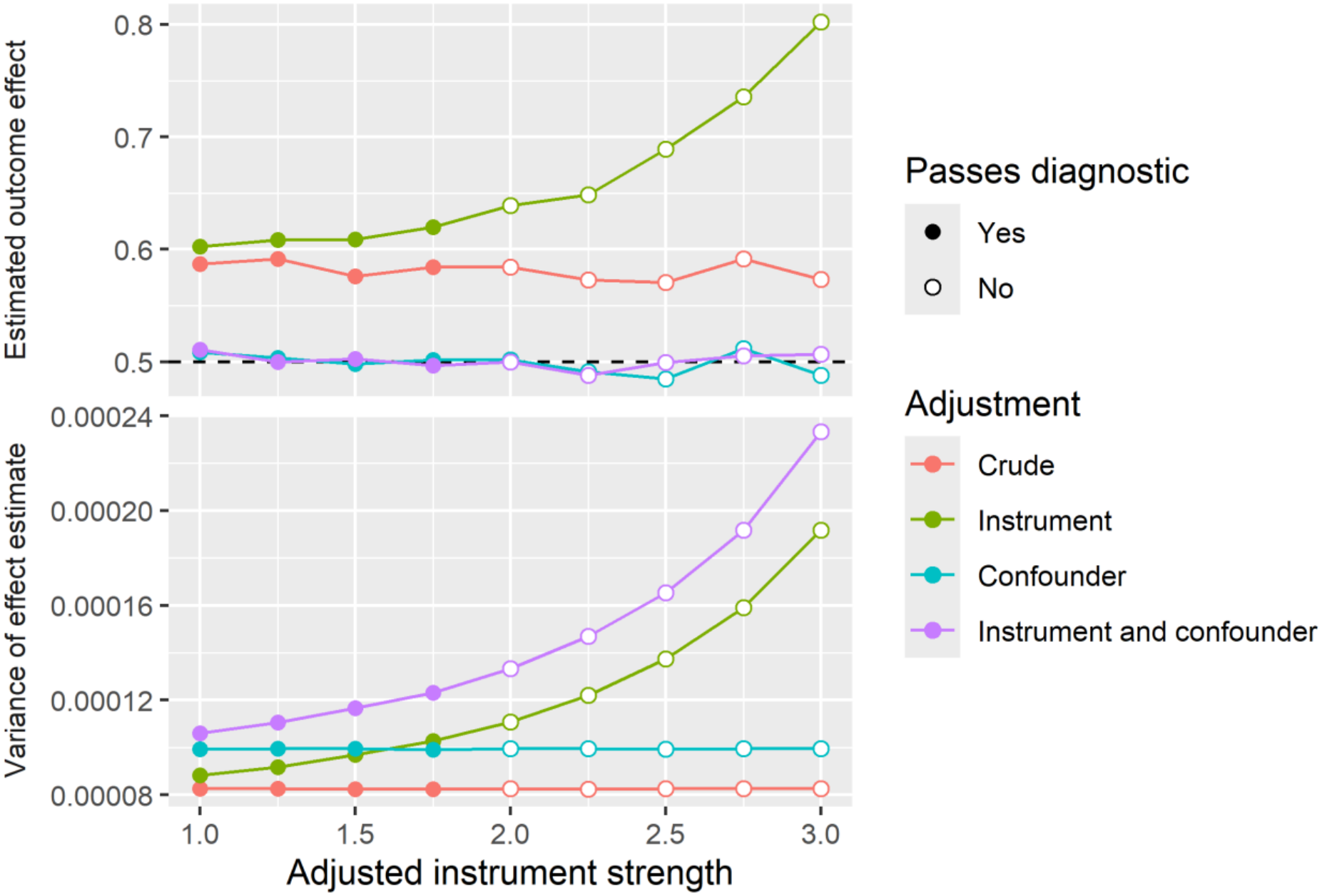
Effect of adjusting for 100 instruments on estimation. The estimated effect of the treatment on the outcome is shown in the top graph with the correct effect shown as a dashed line at 0.5. Four adjustment methods are shown for a range of adjusted instrument strengths. Analyses that pass the LSPS diagnostics are shown as solid discs, and those that fail are shown as open circles. The bottom graph shows the variance of the effect estimate for the employed analyses and adjusted instrument strengths.

**Figure 3.**
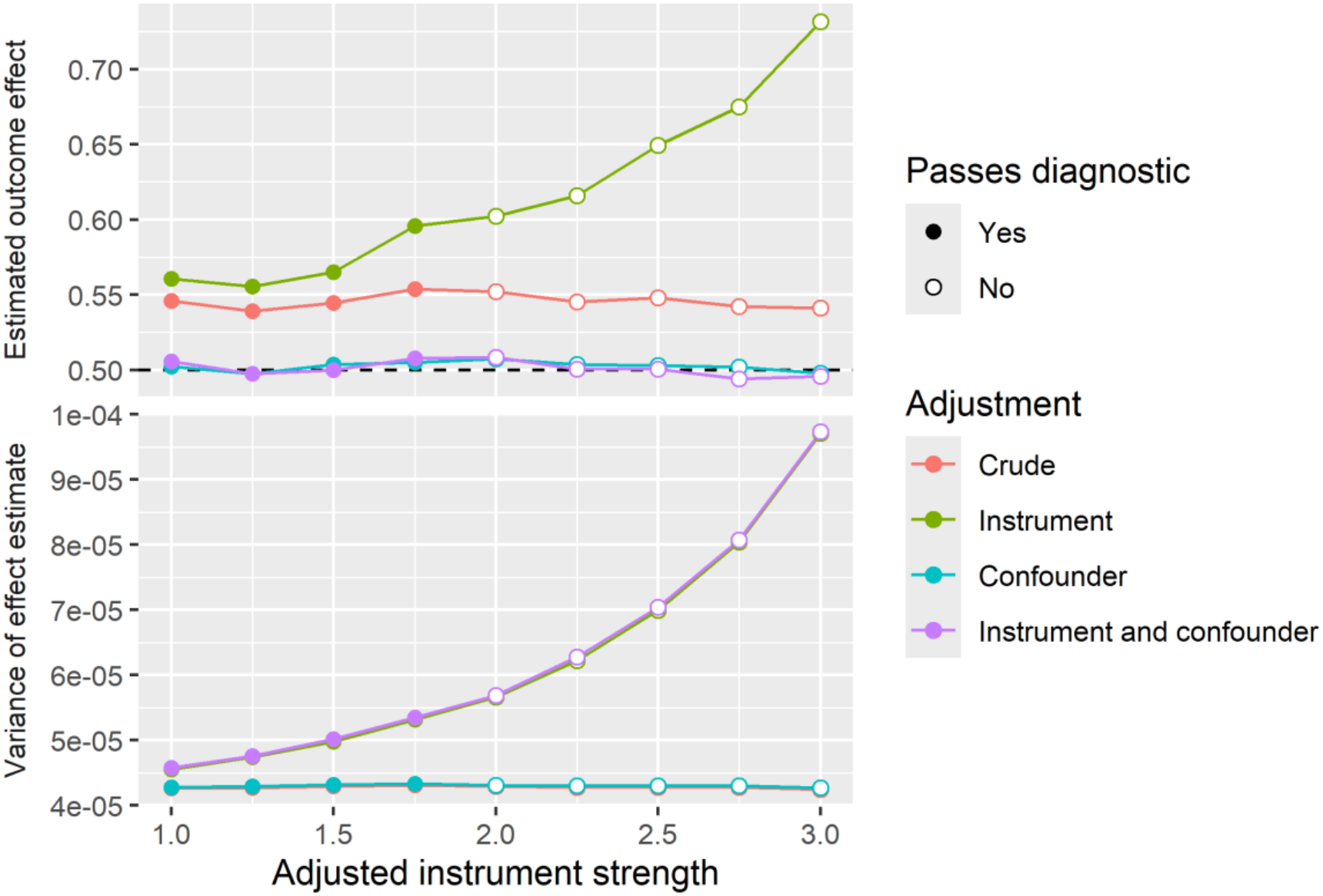
Effect of adjusting for 100 instruments on estimation using a Cox proportional hazards regression on longitudinal time-to-event data. The estimated effect of the treatment on the outcome is shown in the top graph with the correct effect shown as a dashed line at 0.5. Four adjustment methods are shown for a range of adjusted instrument strengths. Analyses that pass the LSPS diagnostics are shown as solid discs, and those that fail are shown as open circles. The bottom graph shows the variance of the effect estimate for the employed analyses and adjusted instrument strengths.

## DISCUSSION

In these simulations, we confirm that adjusting for instruments increases the bias (when there is unmeasured confounding) and variance of the effect estimate. The size of that increase is limited, however, within LSPS’s correlation and equipoise thresholds [22]. Even when the variance of the treatment covariate due to the adjusted instrument is over 20 times greater than the variance due to unadjusted confounding (at *B*=4.65), adjusting for the instrument increases the effect estimate bias by less than that caused by unadjusted confounding. It appears that the quest to adjust for confounding supersedes the risk of adjusting for an instrument if the instrument has a correlation less than 0.5 with the treatment and if equipoise demonstrates that several instruments are not additive.

A threshold on equipoise of 0.5 resulted in minimal bias and minimal excess variance, but the threshold may be overconservative. Equipoise reflects the effect of both adjusted instruments and measured confounders, and only the instrument portion is relevant, so in most studies, equipoise will overestimate the consequence of instruments. With an equipoise threshold of 0.3, excess bias still approximates the bias due to the unmeasured confounding. Conversely, with an equipoise threshold of 0.7, excess bias is minimal.

This simulation aligns with a previous assessment of the impact of adjusting for an instrument [27], which found that simulated instruments had a weak effect on bias and precision in both simulations and negative control experiments based on real-world data. It also aligns with the empirical evidence that adjusting for large numbers of covariates via LSPS produces less biased results than manual selection [23,24] or empirical selection [21] despite the possibility that using large numbers of covariates may include mild to moderate instruments.

Our study is limited in being a simulation. Its purpose is to explore the relationship between induced bias by adjusting for instruments to thresholds used in LSPS. Additional studies using real-world data may be warranted, although some of that work has been done [27].

Our results favor the use of large-scale propensity score adjustment despite stated concerns in the literature that inadvertent inclusion of instruments may increase bias [19] and despite theoretical concerns, for example, that instrument bias is additive whereas confounding bias may cancel out [18]. The tradeoff between missing confounders and including instruments is an empirical one, depending on the prevalence and strength of confounders and instruments.

Given the use of diagnostics like correlation and equipoise, which we address in this study, the use of large numbers of covariates appears to be effective as shown in previous studies of real-world medical data [21,23,24].

## Data Availability

All data produced in the present work are contained in the manuscript

